# Excitatory and Inhibitory neurochemical markers of anxiety in young females

**DOI:** 10.1101/2023.10.03.23296431

**Authors:** Nicola Johnstone, Kathrin Cohen Kadosh

## Abstract

Between the ages of 10-25 years the maturing brain is sensitive to a multitude of changes, including neurochemical variations in metabolites. Of the different metabolites, gamma-aminobutyric acid (GABA) has long been linked neurobiologically to anxiety symptomology, which begins to manifest in adolescence. To prevent persistent anxiety difficulties into adulthood, we need to understand the maturational trajectories of neurochemicals and how these relate to anxiety levels during this sensitive period. We used magnetic resonance spectroscopy in a sample of younger (aged 10-11) and older (aged 18-25) females to estimate GABA and glutamate levels in brain regions linked to emotion regulation processing, as well as a conceptually distinct control region. Within the Bayesian framework, we found that GABA increased, and glutamate decreased with age, negative associations between anxiety and glutamate and GABA ratios in the dorsolateral prefrontal cortex, and a positive relationship of GABA with anxiety levels. The results support the neural over-inhibition hypothesis of anxiety based on GABAergic activity.

---

The neurochemical gamma-aminobutyric acid (GABA) is the primary metabolite responsible for inhibitory processes in the brain. GABA has long been linked neurobiologically to anxiety symptomology (Lydiard, 2003). By consensus, it is typically reasoned that increased levels of GABA in the brain equate to low anxiety. However, recent research suggests that the opposite may be the case, and the inhibitory function of GABA actually relates to higher levels of anxiety because it serves to over-regulate higher-level cognitive function (Page & Coutellier, 2019). This contrasting proposition aligns better with what is known about the cognitive neurobiology of anxiety. Here we present neuroimaging evidence from emotion regulation regions in young females that supports the proposal that greater levels of GABA are related to higher levels of anxiety.

The functional neural networks involved in emotion regulation development are well defined, with interactions between the subcortical limbic regions, including the amygdala and insula, and the prefrontal cortex, including the dorsolateral prefrontal cortex and anterior cingulate cortex (Ahmed et al., 2015). While the subcortical regions are reactive drivers of the emotional experience, the cortical regions temper responses (Mauss et al., 2007). The ability to do so is a result of maturation, contingent on experience, refined throughout adolescence (Thompson et al., 2008). Adolescence has been recognised as an important maturational period in brain development where functional connectivity increases (Fair et al., 2008) and explicit learning of emotional dampening occurs (Etkin et al., 2015). Neurochemicals have a foundational role in developing functional complexity through neurotransmission and modulating synaptic function of neurons (Huang et al., 2007).

Glutamate and GABA are two such neurochemicals, found throughout the central nervous system exhibiting excitation and inhibitory functions respectively (Elliott, 1965). Synapse formation, the basis of early functional cortical development, is underpinned by GABA led activation stimulating a glutamate response that shapes functional connectivity (Ben-Ari, 2002; Owens & Kriegstein, 2002). While early development is characterised by critical periods triggered by the maturation of localised neurocircuitry involving GABA (e.g., in the visual cortex (Levelt & Hübener, 2012) and in speech perception (Werker & Hensch, 2015)), others have postulated that a reduction in the ratio of Glutamate and GABA, or of excitation to inhibition (E/I), is revealing of sensitive periods in adolescence (Larsen et al., 2022). Thus, prolonged neuronal adaptations, as seen in the developing human brain, are supported by malleable glutamate and GABA activities in functional networks. For example, during mid-childhood and adolescence glutamate and GABA have been implicated in neuroplastic processes supporting cognition (Cohen Kadosh et al., 2015; Stanley et al., 2017; Zacharopoulos et al., 2021). The plasticity in neural circuitry during the period from 10-25 years coincides with the onset of the majority of mental health disorders, including anxiety. Thus, finding therapeutic targets, such as neurochemical markers, to prevent persistent mental health difficulties is a matter of priority (Uhlhaas et al., 2023).

Anxiety aetiology is complex, but certainly founded in neurobiological processes within which GABA and glutamate have critical roles (Millan, 2003). In computational neuroscience, functional stability in operational brain circuitry is described as an attractor network (Khona & Fiete, 2022), a sub-system of cooperating units in a dynamic system. The neural circuitry of anxiety has been described in a dynamic systems network of feedback loops that hinge on the balance of glutamate and GABA neurotransmitters (LeDuke et al., 2023). It has been proposed that anxious states are reached and maintained by the tipping of a stable attractor network towards dominance of bottom-up processing (e.g., sensory driven) over top-down processing (e.g., cognitively evaluated) (LeDuke et al., 2023). Herein, the attractor network and anxiety neurocircuitry are referenced by excitability, or glutamate activity, whereby over-arousal states are construed as under-regulated (e.g., lacking inhibitory balance) in the bottom-up network.

Alterations in the attractor networks describing the neurocircuitry of anxiety and depression implicate changes to the balance of glutamate and GABA. Several empirical studies have queried the role of glutamate and GABA in in anxiety and depression using ^1^H-MRS of the human brain, focused on the prefrontal cortex. While some found positive correlations of GABA and anxiety (Delli Pizzi et al., 2016), others found evidence for the opposite negative correlation (Hasler et al., 2009, 2019; Long et al., 2013). The purpose of this study was to evaluate the relationship of glutamate and GABA with measures of anxiety in two healthy, typically developing female samples. Using ^1^H-MRS in regions of the brain related to emotion regulation against a control region, we assessed whether and how neurochemical levels were correlated in the period between 10-25 years old, and to determine the relationship with pre-clinical anxiety levels young females. We hypothesised that glutamate and GABA would be related to age, and to levels of trait anxiety, social anxiety, and depression, with a particular role for the DLPFC, a key region in emotion regulation.

## Methods

### Participants

The sample size was not predetermined. Data was collected from two independent studies using the same protocols in young females. There was a younger group; *n* = 49, aged 10-12 years (*M* = 132 months), and an older group; *n* = 32, aged 18-25 years (*M* = 260 months). For individuals under 16 years, primary caregivers provided written informed consent as parents/guardians, and child participants provided assent prior to study participation. Individuals over 16 years supplied written consent themselves. All participants were neurologically healthy with no past or current psychiatric diagnosis, neurodevelopmental condition, or mental health difficulties. Participants were compensated for their time (£25/hour). The local Research Ethics committee reviewed data collection protocols and provided favorable ethical opinion.

### Procedure

Both samples completed age appropriate questionnaires on trait anxiety, social anxiety, and depression (detailed below) prior to having a brain scan. Brain scans consisted of obtaining images for constructing the anatomical structure so that the volumes of interest (VOIs) for magnetic resonance spectroscopy imaging could be located. In the younger group three VOIs were defined over the left amygdala, left dorsolateral prefrontal cortex and the left inferior occipital gurus. In the older group, the same regions were used except the amygdala was replaced with a region over the anterior cingulate cortex. Brain scans lasted approximately 1 hour in both groups.

### Questionnaires

#### Trait anxiety

The State-Trait Anxiety Inventory for Children (STAI-C, Spielberger, 1973) was used for the younger group. This consists of two 20-item scales that measure state and trait anxiety in children between the ages of 8 and 14, with separate scores for each. Each scale gives a statement and 3 phrase responses of which one is selected. On the trait scale respondents are asked to consider how they generally feel, with options hardly = 1, sometimes = 2, often = 3. Scores range from 20-60. Similarly, the older group completed the State-Trait Anxiety Inventory (STAI, Spielberger et al., 1983),again with 20 questions each for state and trait anxiety scales. All items are rated on a 4-point (1-4) scale (e.g., from “Almost Never” to “Almost Always”), with a range on the trait scale from 20 – 80. For each questionnaire, higher scores indicate higher trait anxiety levels. Reliability for each sample was good (20 items, younger α = .80, older α = 0.93).

#### Social anxiety

Both younger and older groups were given the Social Anxiety Scale for Children-Revised (SAS, La Greca, 1999). A 22 item self-report measure of 3 factors related to social anxiety; fear of negative evaluation (8 items); social avoidance and distress in new situations (6 items) and social avoidance and distress in general (4 items). Items are rated on a 5 point Likert scale (1 = not at all, 2 = hardly ever, 3 = sometimes, 4 = most of the time, 5 = all the time). Each subscale is summed to provide a total social anxiety score used here, ranging between 18-90. Higher values equate to more social anxiety. Reliability for each sample was good (18 items, younger α = .69, older α = 0.69).

#### Depression

Child Depression Inventory (CDI, Kovacs, 1992) is a 27-item rating instrument with groups of three sentences to choose from, and requests the selection of one based on how they have been feeling over the past two weeks. Sentences are scored a = 0; b = 1; c = 2 and totals are summed. Similarly, the older group were given the Beck Depression Inventory (BDI-II, Beck et al., 1996). This is a 21 item self-report using a four-point scale ranging from 0 (symptom not present) to 3 (symptom very intense). Values are summed, greater values indicate more depression symptoms. Reliability for each sample was good (27 items, younger α = .80; 21 items, older α = 0.87).

### MRI acquisition

For both groups, MRI data were acquired on a Siemens 3T Magneton TIM Trio scanner with a 32-channel head coil. Sagittal T1-weighted Magnetization Prepared Rapid Acquisition Gradient Echo (MPRAGE) images were acquired under the parameters TR = 1900 ms, TE = 3030 and dwell time 1500 ms in 1 mm slices in field of view (FOV) resolution 256 x 256 over 5 minutes acquisition time. T2*-weighted Turbo Spin Echo (TSE) images were acquired on the coronal and axial planes with TR = 3500 ms, acceleration factor (GRAPPA = 2), TE = 93 ms and dwell time 7100 ms in FOV of 179 x 256 in 4 mm slices. These structural images were then reconstructed for planning voxel placement for ^1^H-MRS acquisition.

### ^1^H-MRS data acquisition

^1^H-MRS in vivo spectra were acquired in three volumes of interest (VOIs), for each group. In the older group the VOIs were the left dorsolateral prefrontal cortex (DLPFC), anterior cingulate cortex (ACC) and left inferior occipital gyrus (IOG) using a 2 cm^3^ voxel manually centred in reference to the sagittal acquisition over the VOI. In the younger group, the DLPFC and IOG VOIs were similarly obtained in addition to a VOI over the left amygdalae using a 1.5 cm^3^ voxel.

Spectra were acquired using the Spin Echo full Intensity-Acquired Localized spectroscopy sequence, (SPECIAL, Mlynárik et al., 2006). Water suppressed spectra were acquired with TR = 3200 ms, TE 8500 ms, flip angle of 90° over 192 averages (245 averages for the amygdala VOI). Immediately prior, water unsuppressed spectra were acquired over 16 averages in the same region. Saturation bands were manually aligned along the six outer planes of each voxel prior to acquisition to suppress undesirable signals beyond the voxel boundaries.

### ^1^H-MRS data pre-processing

For each VOI, motion corrupted averages and frequency drift were corrected using processing functions in the FID-A toolbox (www.github.com/CIC-methods/FID-A, Simpson et al., 2017) in MATLAB (The Mathworks, Natick, MA). Cleaned averaged spectra were then analysed in LCModel (Provencher, 1993) to estimate relative concentrations of GABA and glutamate in each VOI in reference to the water peak of each VOI. Four measures of spectral quality were obtained to determine adequate quality for inclusion in analysis. 1. The line width of the water peak (full width at half maximum height) with widths greater than 13 Hz excluded. 2. The signal to noise ratio (SNR, signal [height of NAA peak found between 1.8-2.2 ppm] divided by noise [averaged signal between the metabolite free range −2 ppm – 0 ppm]). Plots of the SNR distributions in each VOI for each group determined a SNRs less than 150 to be poorly estimated and excluded. 3. Visual inspection corroborating the visibility of the NAA, Cr, Cho, and Glutamate peaks at the expected ppm, and 4. Cramer-Rao lower bounds (CRLBs) of each metabolite estimated. Higher CRLBs are typically coupled with lower metabolite estimates however in the case of low volume metabolites such as GABA it is difficult to determine if the CRLBs estimates are induced by low levels or poor signals. Considering our stringent quality inspection methods, and additional plotting and inspecting of the CRLBs against the estimated metabolites, CRLBs were determined for each metabolite separately, but were consistent across younger and older groups, and VOIs. For GABA, estimates with CRLBs>29 were excluded, and for glutamate, estimates with CRLBs>15 were excluded. Outcomes of the inspection protocol are displayed in **Table 1**. Note, none of the amygdala recordings were of suitable quality for further analysis.

**Table 1.** Overview of metrics applied to assess signal quality of spectra acquired in each VOI for younger and older age groups, and number of surviving datasets.

| Characteristic | Older |  |  | Younger |  |  |
| --- | --- | --- | --- | --- | --- | --- |
|  | ACC, N = 32 | DLPFC, N = 32 | IOG, N = 32 | AMY, N = 49 | DLPFC, N = 49 | IOG, N = 49 |
| SNR mean (SD) | 308 (54) | 228 (60) | 264 (49) | 158 (NA) | 280 (51) | 258 (67) |
| N < 150 | 0 | 0 | 1 | 48 | 9 | 10 |
| FWHM (Hz) mean (SD) | 6.57 (1.26) | 7.27 (0.62) | 7.58 (0.90) | 10.29 (2.11) | 6.50 (0.47) | 7.60 (1.02) |
| N > 13 Hz | 0 | 0 | 1 | 29 | 9 | 8 |
| CRLB: GABA mean (SD) | 11.3 (2.7) | 12.7 (2.8) | 12.8 (3.7) | 16.6 (4.5) | 12.9 (3.2) | 14.0 (4.7) |
| N > 29 | 1 | 3 | 1 | 30 | 3 | 4 |
| CRLB: Glutamate mean (SD) | 2.88 (0.71) | 3.91 (1.53) | 4.22 (1.21) | 8.14 (2.68) | 3.35 (1.98) | 4.36 (1.73) |
| N > 15 | 0 | 0 | 0 | 20 | 1 | 4 |
| Surviving datasets | **31** | **29** | **30** | **0** | **39** | **38** |

*Note.* SNR = signal to noise ratio. FWHM = full width half maximum. CRLB = Cramer-Rao lower bounds.

Voxel based morphometry was performed in the MATLAB Statistical Parametric Mapping toolbox (SPM12) to estimate grey matter (GM), white matter (WM) and cerebrospinal fluid (CSF) composition of each VOI. Corrected metabolite concentrations were calculated on the assumption that there is twice as much metabolite concentration in grey matter as white matter, and negligible in cerebral spinal fluid, thus followed the formula:

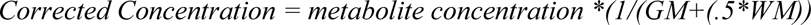

### Statistical analyses

This exploratory analysis aimed to uncover if there is a relationship between anxiety levels in typically developing healthy females and levels of GABA and glutamate in brain regions implicated in emotion processing. Correlational analyses were used to determine if a relationship exists, and the direction of effects. We opted to perform correlational tests under a Bayesian framework which is more powerful in quantifying true effects where prior knowledge is uncertain.

Descriptive point estimates of the questionnaires, and neurochemical levels are provided as means, and variance quantified as standard deviations for the younger and older groups separately. For the correlations, we first examined if there was a relationship with age and anxiety, depression, and social anxiety for the younger and older groups separately. We then examined if there was a relationship between age and grey matter, and glutamate and GABA. Outcomes were then standardised for each group independently to set younger and older groups on an equivalent scale, and the relationships between anxiety, depression and social anxiety, and the neurochemicals GABA and glutamate examined using Bayesian Pearson correlations of rank transformed data. The point estimate Rho is reported with 95% credible interval, the probability of effect direction (pd, >95% indicative of effect) and region of practical equivalence (ROPE, defined −0.05, 0.05, < 2.5% in ROPE indicative of significant effect), with the Bayes factor (BF). Note BF_10_ denotes evidence in favour of the alternative hypotheses. Evidence is quantified as anecdotal (BF_10_ 1-3), moderate (BF_10_ 3-10), strong (BF_10_ 10-30), very strong (BF_10_ 30-100) and extreme (BF_10_ >100) (Lee & Wagenmakers, 2014). Analyses were performed in R version 4.3.0 (R Core Team, 2023) using the correlation package (Makowski et al., 2020, 2023)

## Results

### Descriptive statistics

Of 32 females in the older group, and 49 in the younger group, 32 and 47 respectively provided responses to the questionnaires. For all measures mean scores were aligned with the norms for each age group (**Table 2**). **Table 2** also presents the means and standard deviations of the measures obtained in each VOI in the brain from the younger and older age groups.

**Table 2.** Means and standard deviations of measures obtained in each VOI in the brain from the younger and older groups.

|  |  | Older (18-25 years) |  |  | Younger (10-11 years) |  |
| --- | --- | --- | --- | --- | --- | --- |
| Questionnaires |  | N = 32<sup>1</sup> |  |  | N = 47<sup>1</sup> |  |
| Trait anxiety |  | 44.39 (9.71) |  |  | 32.02 (7.87) |  |
| Social anxiety |  | 51.29 (11.02) |  |  | 43.70 (11.32) |  |
| Depression |  | 12.29 (9.06) |  |  | 6.77 (6.07) |  |
| Neuroimaging |  | ACC, N = 31<sup>1</sup> | DLPFC, N = 29<sup>1</sup> | IOG, N = 30<sup>1</sup> | DLPFC, N = 39<sup>1</sup> | IOG, N = 38<sup>1</sup> |
| Grey matter (%) |  | 67 (4) | 35 (7) | 37 (5) | 41 (6) | 47 (7) |
| White matter (%) |  | 14 (3) | 64 (7) | 62 (5) | 58 (7) | 53 (7) |
| GABA (i.u) |  | 1.914 (0.589) | 2.202 (0.646) | 2.374 (0.643) | 1.792 (0.424) | 2.251 (0.764) |
| Glutamate (i.u) |  | 8.806 (0.803) | 8.671 (1.057) | 7.609 (1.154) | 9.348 (0.482) | 8.366 (1.022) |
| Ratio Glutamate: GABA |  | 4.885 (1.124) | 4.279 (1.330) | 3.470 (1.188) | 5.472 (1.196) | 4.072 (1.367) |

<sup>1</sup>Mean (SD)*Association of VOI measures with age.*

### Associations of questionnaire responses with age

Bayesian Pearson Correlations were conducted between age (in months) and trait anxiety, depression, and social anxiety for the younger and older groups separately. There was no evidence for any associations of age in months with social anxiety, trait anxiety, or depression (**Table S1**).

### Association of VOI measures with age

Bayesian Pearson Correlations were conducted between age (in months) and measures obtained from each VOI (grey matter, GABA, Glutamate and the ratio of glutamate and GABA). In each the DLPFC and IOG there was extreme evidence for a reduction of grey matter with increasing age in months (DLPFC, *rho* (66) = −0.442, 95% CI[−0.618, −0.225], pd = 100%, *ROPE* = .08%, BF_10_ >100; IOG, *rho* (66) = −0.575, 95% CI[−0.717, −0.406], pd = 100%, *ROPE* = 0%, BF_10_ >100). Additionally, there was anecdotal evidence of the same effect in the ACC, *rho* (30) = −0.238, 95% CI[−0.570, −0.015], pd = 95.9%, BF_10_ = 1.671, which was recorded in the older group only (aged 18-25 years). These effects are displayed in **Figure 1A**. The DLPFC further showed extreme evidence that ratios reduce with increasing age, *rho* (66) = −0.417, 95% CI[−0.617, −0.222], pd = 99.8%, *ROPE* = 0.35%, BF_10_ >100, accompanied by very strong evidence of increasing GABA levels with age in months, *rho* (66) = 0.387, 95% CI[0.165, 0.564], pd = 99.98, *ROPE* = 0.53%, BF_10_ = 72.762, and very strong evidence of reducing glutamate levels with age in months, *rho* (66) = −0.357, 95% CI[−0.543, −0.147], pd = 99.95%, *ROPE* = 0.73%, BF_10_ = 31.622. Like the DLPFC, the IOG showed strong evidence for a reduction in glutamate levels with age in months and anecdotal evidence for a negative association with ratios and increasing age in months. There was no evidence relating age in months to neurochemicals in the ACC (**Table S2**). This is presented in **Figure 1B**.

**Figure 1.**
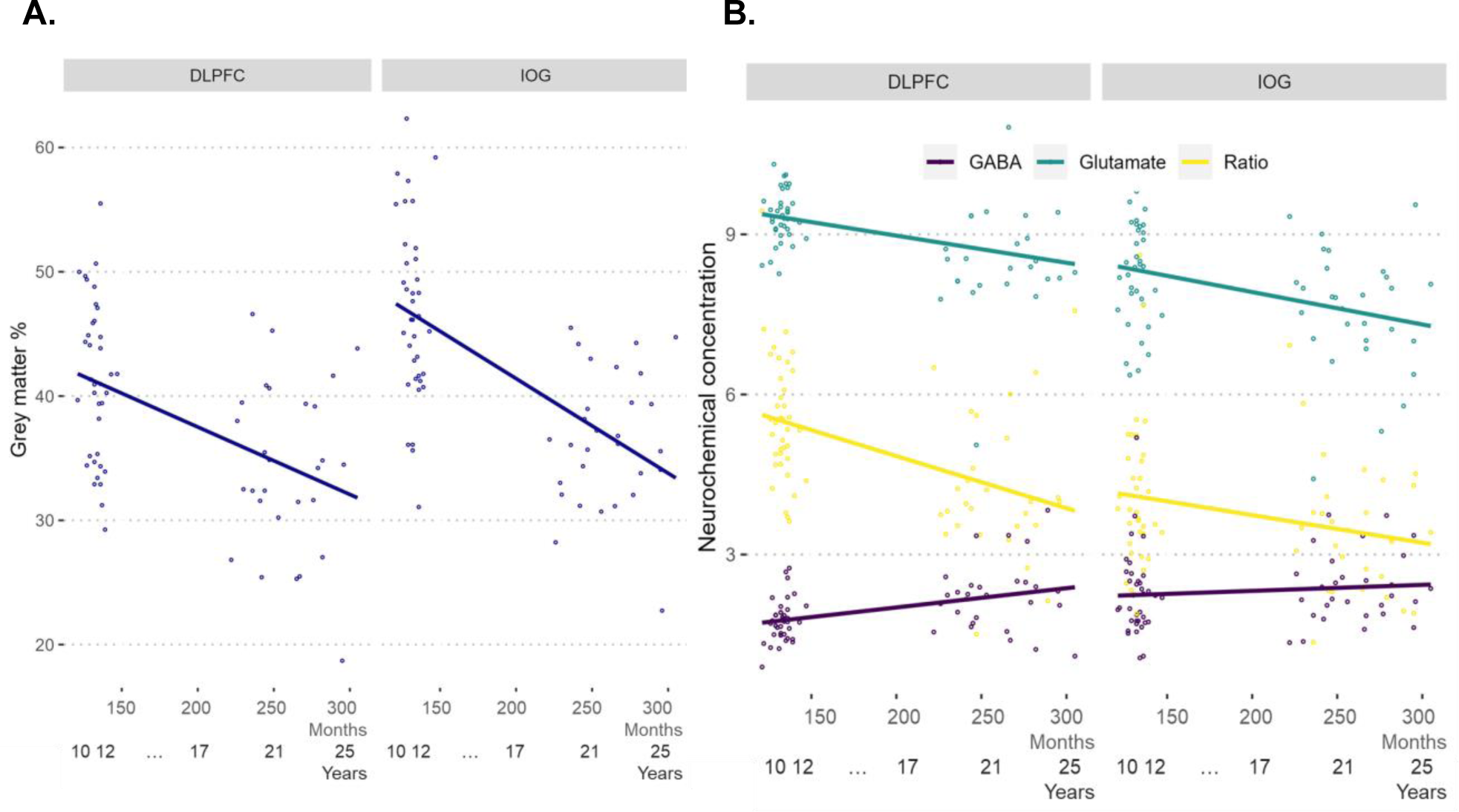
Panel A plots the negative realtionship between grey matter volume and increasing age in months, this was extreme and significant the DLPFC and IOG, and anecdotal in the ACC (not shown). Panel B illustrates the association between the neurochemicals GABA (in purple), glutamate (in green), and the ratio (in yellow) of these against increasing age in months. Effects were most prominent in the DLPFC, where there was extreme evidence for a significant negative association between ratios and age in months, very strong evidence of a positive realtionship between GABA and age in months, and very strong evidence for a negative relationship of gluatmate and increasing age. The IOG showed strong evidence for a significant neagtive realtionship of glutamate with age in months, and anecdotal evidence of a negative association of ratios with age.

### Association of neurochemicals with questionnaire responses

The evaluation of evidence for associations between questionnaire responses and the neurochemicals in each VOI was also conducted using Bayseian Pearson correlations. As predicted, there was more quantifiable evidence for the role of neurochmicals in the DLPFC in relation to anxiety measures compared to the IOG, or ACC. In the DLPFC, the ratio of glutamate to GABA had a moderate negative assoication with social anxiety, *rho* (66) = −0.286, 95% CI[−0.472, −0.064], pd = 99.20%, BF_10_ = 5.758, and trait anxiety, *rho* (66) = −0.263, 95% CI[−0.473, −0.057], pd = 98.98%, BF_10_ = 3.355, illustrating that larger ratios are related to lower levels of anxiety. In the DLPFC, GABA was anecdotally shown to have a positive relationship with social anxiety, *rho* (66) = 0.207, 95% CI[−0.032, 0.415], pd = 95.63%, BF_10_ = 1.336 and trait anxiety, *rho* (66) = 0.223, 95% CI[0.011, 0.459], pd = 96.70%, BF_10_ = 1.743, while glutamate was anecdotally negatively associated with social anxiety only, *rho* (66) = −0.193, 95% CI[−0.398, 0.035], pd = 95.63%, BF_10_ = 1.142. Interestingly, GABA was anecdotally positively related to social anxiety in the IOG, *rho* (66) = 0.219, 95% CI[−0.005, 0.432], pd = 97.28%, BF_10_ = 1.670. There was no evidence of a relationship in any VOI to depression. There full results are presented in **Table S3** and plotted in **Figure 2**.

**Figure 2.**
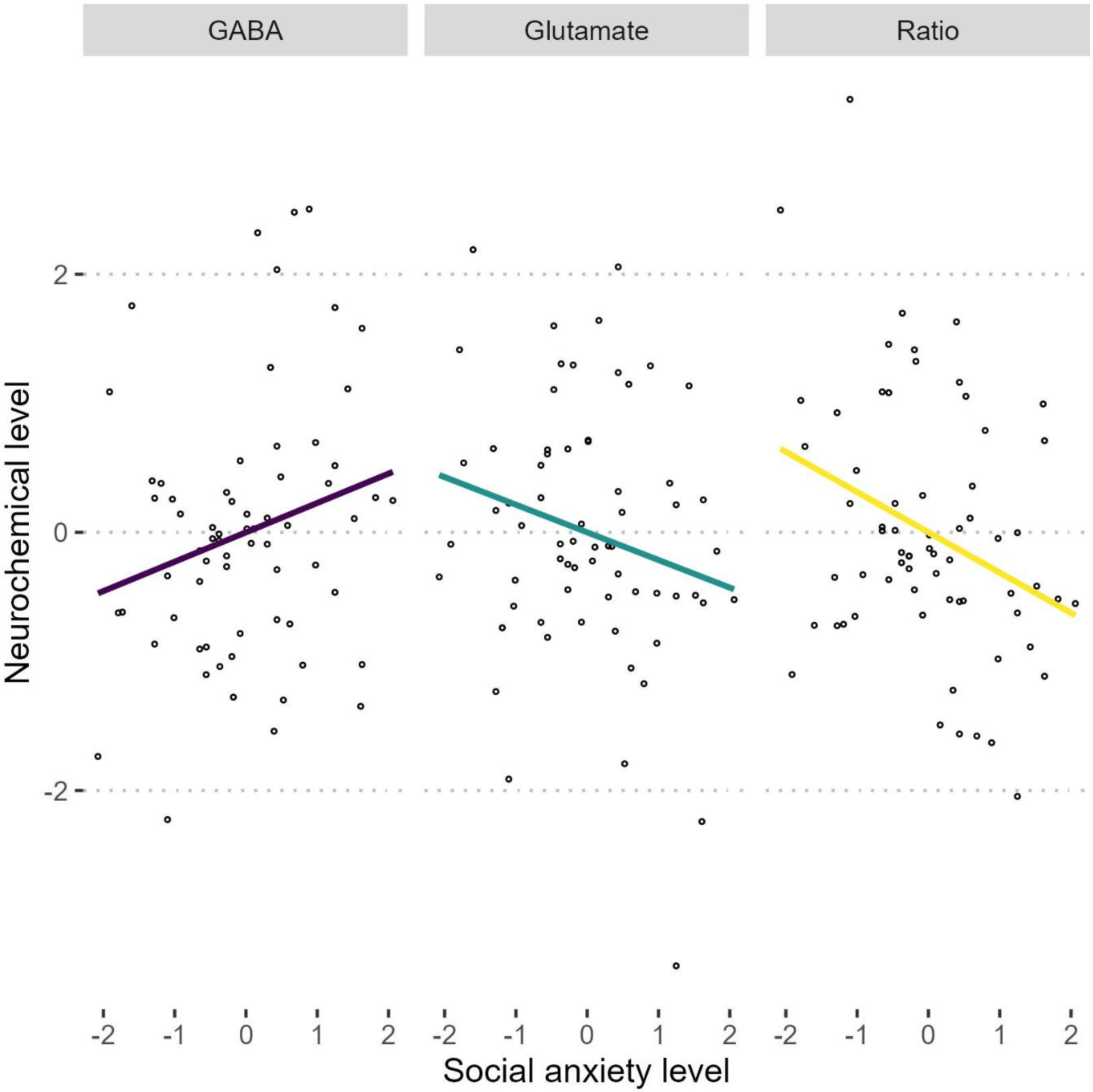
Plot of the neurochemical associations in the DLPFC against social anxiety. The neurochemical GABA is plotted in purple, glutamate in green, and ratios in yellow. Social anxiety was found to have a negative moderate relationship with glutamate and GABA ratios, a negative relationship with glutamate and a positive relationship with GABA in both the DLPFC and IOG (not shown). Trait anxiety also showed a modereate negative relationship with ratios, similar to social anxiety.

## Discussion

We found evidence for inhibitory and excitatory neurochemical markers of anxiety in young females, supporting our hypothesis that the DLPFC is a key region in linking neurochemical levels to experiences of anxiety. We determined that that the balance of glutamate and GABA is negatively related to increasing levels of social and trait anxiety, suggesting that smaller ratios are related to greater anxiety. This was coupled with evidence suggesting a positive relationship of anxiety with GABA, and negative relationship of anxiety with glutamate. Thus, greater levels of inhibitory control are associated with more anxieties.

Despite past inconsistency in findings of the role of GABA in stress, anxiety, and depression, evidence is emerging to suggest that over-inhibition of function in the prefrontal cortex, driven by the over representation of GABA in reference to glutamate might be key in understanding the neural circuitry of anxiety. In a reconciliation of these inconsistent findings, a recent review proposes that hypoactivity in the prefrontal cortex induced by stressors preceding emotional processing difficulties result in an imbalance in glutamate and GABA functioning (Page & Coutellier, 2019). Critically, the principle of homeostatic plasticity would allow GABA/glutamate to adaptively respond by increasing or reducing activity as required. However, in states where excitatory function is reduced, inhibitory function will be over represented (Page & Coutellier, 2019). In this way, *over-inhibition* of prefrontal function observed in anxiety and depression may be driven by relative increases of GABAergic activity in relation to reduced glutamatergic activity.

Our data support this conceptualisation, although we have measured concentrations of neurochemicals rather than activity or function produced by the neurochemicals. Yet, we assume that circulating concentration levels measured by ^1^H-MRS are representative of activity, as has been found to be true in animal models (Takado et al., 2022). Thus, here we show lower ratios of glutamate and GABA are related to higher levels of anxiety, driven by higher levels of GABA (relative to glutamate). Note, GABA driven over-inhibition in the PFC has also been observed during cognitive tasks, leading to the proposal that this reduces cognitive flexibility in higher level information processing (Li et al., 2022). In one step further, evidence from animal models suggests that higher levels of tonic GABA can be disruptive to higher order cognitive functions, and facilitate processing in sensory regions (Koh et al., 2023).

Assembling this evidence, we support the conceptualisation of the neurocircuitry of anxious states as an imbalanced attractor network referenced by glutamatergic activity as proposed by LeDuke and colleagues (2023), whereby over-inhibition may boost sensory processing and/or impair higher order cognition, with the caveat that thorough investigation of the neurochemistry of different nodes involved in top-down and bottom-up processing is required. For now, we conceive our findings to represent a neural over-inhibition hypothesis of anxiety. Imagined as a net in which threads of GABA and glutamate are weaved together, with the balance, or ratio representing the density of the net. A less dense net allows functional free flow of information. Where GABA increases relative to glutamate and the density of the net increases e.g., the ratio becomes smaller, resulting in smaller holes that do not allow easy diffusion of information, thus signals become trapped, propagating anxious states. We propose that higher levels of GABA, as measured by ^1^H-MRS will be related to higher levels of anxiety, representing functional over-inhibition.

These data also showed that with increasing age, there was extreme evidence for negative relationship with the ratio, and very strong evidence for a positive relationship with GABA, and for a negative relationship with Glutamate in the DLPFC, at the same time, there was no evidence of a relationship between age and anxiety levels. Interestingly, in the IOG, a negative relationship of glutamate with age was observed, and a positive relationship found with GABA. This highlights a particular role for GABA in the DLPFC, a region important in emotion regulation processes. However, the decrease of glutamate over time is not region specific, which is indicative of a maturational function of glutamatergic activity, where excitability naturally decreases over time. This maturational decrease in glutamate was also observed in a study of young people age 10-30 years, where glutamate decreased with age (Perica et al., 2022). In contrast to our findings, the balance of glutamate and GABA in the PFC increased through adolescence, where we found the opposite to be true. Interestingly, they also found GABA levels to be stable in the DLPFC, and decrease slightly over time in the ACC. In a large meta-analysis of GABA across the lifespan, it was found that following an early and rapid acceleration in GABA levels during childhood, these begin plateau in the early 20s before slowly declining across the lifespan (Porges et al., 2021). Our data concurs, where we observed a moderate positive increase with age. Note, each of these studies are of cross-sectional design and understanding the maturational trajectory of glutamate and GABA would benefit from longitudinal study. What is clear is that observed concentrations of GABA and glutamate relate to the developmental period of adolescence and are important in maturational outcomes. With anxiety onset originating in mid-childhood, considering GABAergic activity during this period could be important in preventing persistent maladaptive neural function into adulthood.

Our single sex sample contributes to a body of literature suggestive of sex based differences in the neurocircuitry of anxiety. Cohen and colleagues (2023) found that GABA levels in PFC related to different stress neurocircuitry in males compared to females, where males effectively performed better with increased levels of GABA, despite showing no difference in anxiety response or physiologically to stress compared to females (Cohen et al., 2023). This is contrary to the concept that higher GABA levels interferes with cognitive flexibility but might indicate differing thresholds of inhibitory activity in males compared to females. However, in a single sex study of young adult males at 7T, GABA was negatively related to perceived stress in the nucleus accumbens, and positively to the glutamate and GABA ratio (Strasser et al., 2019), contrary to what might be expected in the PFC. This study also measured trait and social anxiety levels and found no relationship to neurochemistry in the nucleus accumbens.

In conclusion, we propose that in developing females presenting with anxiety, lowering brain GABA levels may be a critical preventative step in helping offset maladaptive trajectories into adulthood. However, it is imperative to test our hypothesis that higher GABA levels drive over-inhibition in anxiety in clinical populations and uncover how this interacts with the anxiolytic properties of GABA agonists. Further, the maturational reduction in glutamate levels might be a key anchor of the GABA response, and environmental stimuli may play on this to tip GABA into over-inhibitory states. Improving understanding of the neurocircuitry of anxiety and teasing out what is a typical maturational trajectory and what propagates anxious states will help clinicians determine appropriate therapeutic pathways.

## Data Availability

The data that support the findings of this study are openly available in osf

https://osf.io/thvfe/?view_only=50b8d0a854be42968e7d5efc982902f3

## Acknowledgements

This study was supported by an Academy of Medical Sciences/Wellcome Trust Springboard fellowship to K.C.K. (HOP001\1004). We thank Phoebe Crane, Oliver Crenol, Rothaa Guidarelli Mattioli, Hannah Piggott, Annalisa Schliephake, Hannah Baron, Susannah Dart and Chrzanowski for help with data collection and pre-processing, and Professor Harriet Tenenbaum for project support and comments on an earlier version of the manuscript.

## Conflict of interest statement

The authors declare no interest or relationship, financial or otherwise as influential to the work presented here.

## Author note: data sharing

The data that support the findings of this study are openly available in osf at https://osf.io/thvfe/?view_only=50b8d0a854be42968e7d5efc982902f3

---

**Table S1.**
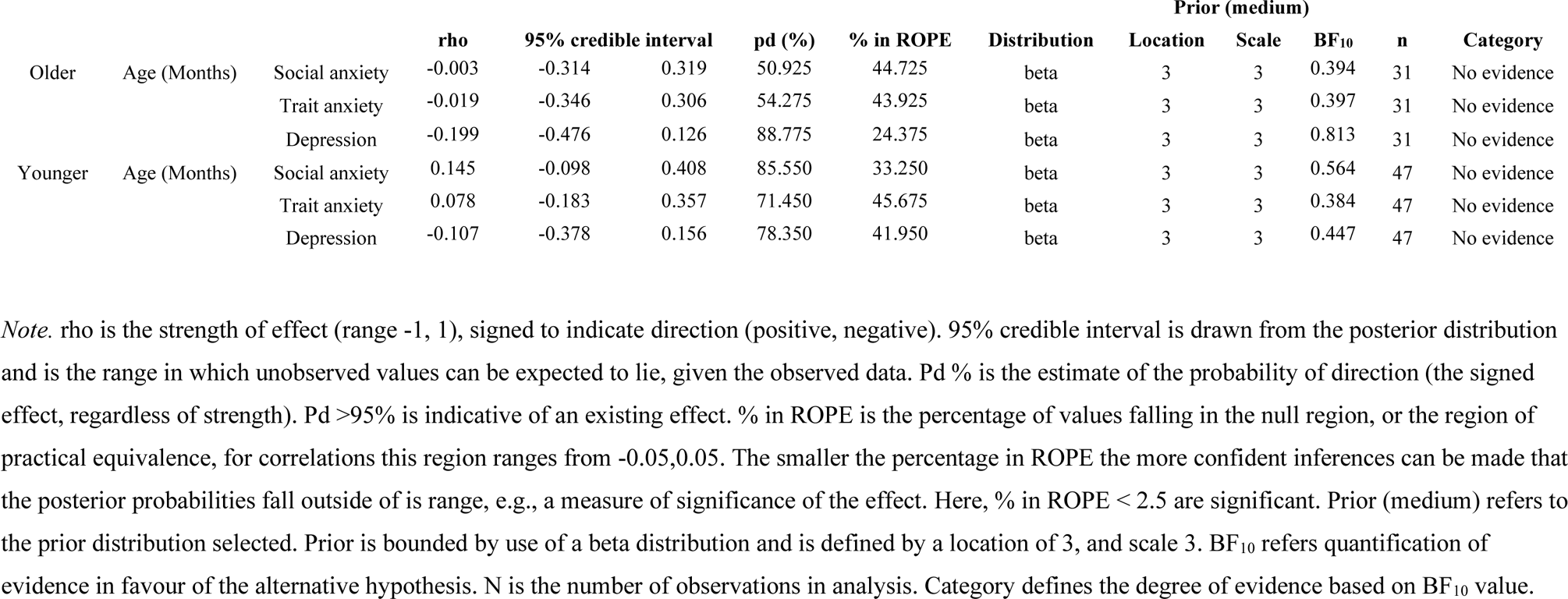
Bayesian Pearson Correlations between age in months and each measure of questionnaire responses for the younger and older age groups.

**Table S2.**
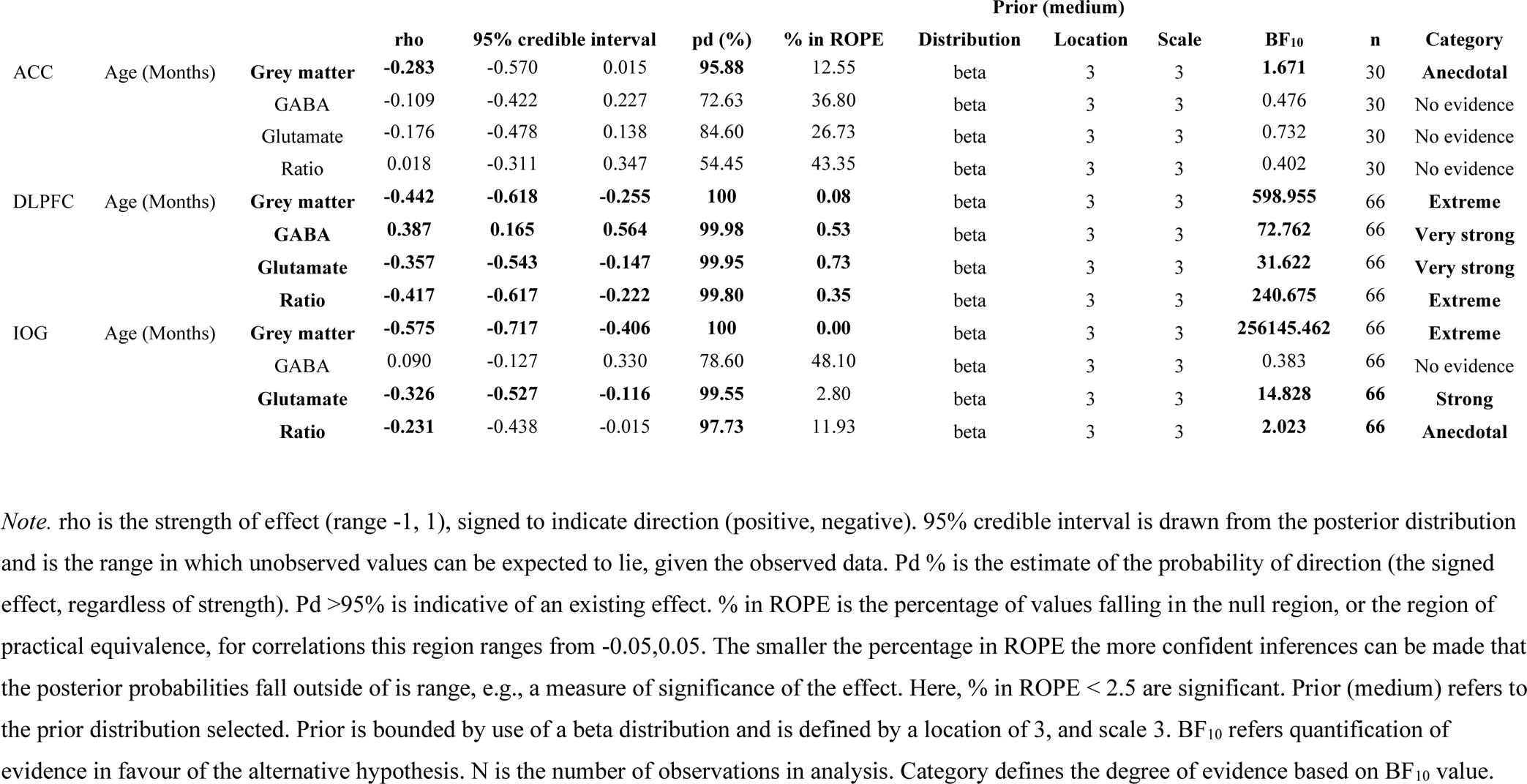
Bayesian Pearson Correlations between age in months and each neurochemical in each region.

**Table S3.**
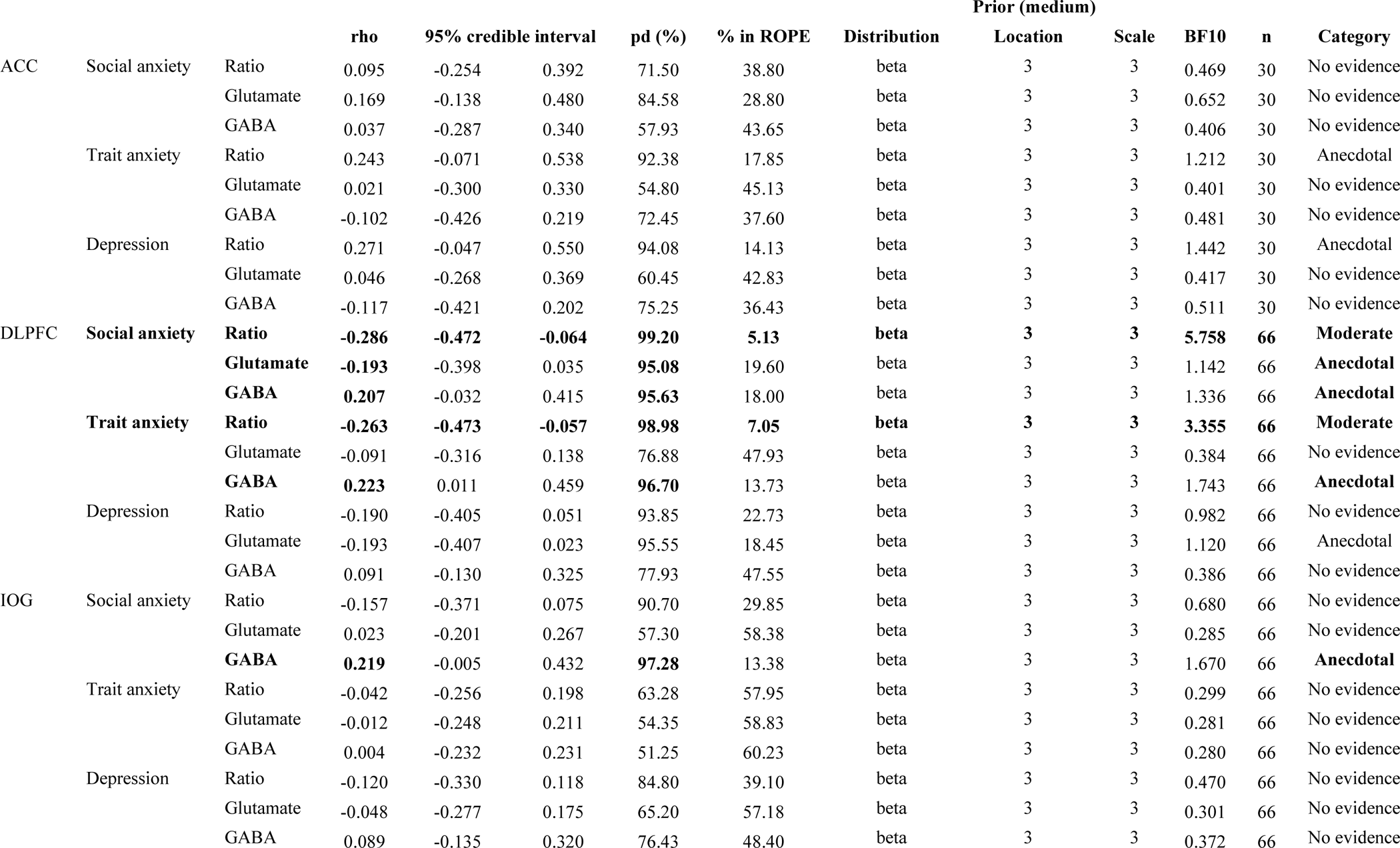
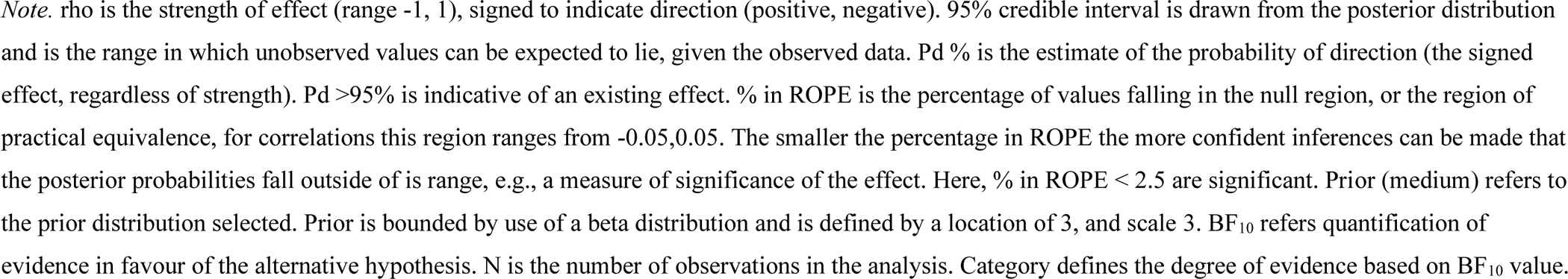
Bayesian Pearson Correlations between measures of questionnaire responses and neurochemicals in each region, standardised by sample.

## Notes

### Competing Interest Statement

The authors have declared no competing interest.

### Author Declarations

The University Ethics Committee of the University of Surrey gave ethical approval for this work.

## References

Ahmed, S. P., Bittencourt-Hewitt, A., & Sebastian, C. L. (2015). Neurocognitive bases of emotion regulation development in adolescence. Developmental Cognitive Neuroscience, 15, 11–25. 10.1016/j.dcn.2015.07.006

Beck, A. T., Steer, R. A., & Brown, G. K. (1996). BDI-II: Beck Depression Inventory Manual. (2nd ed.). Psychological Corporation.

Ben-Ari, Y. (2002). Excitatory actions of gaba during development: The nature of the nurture. Nature Reviews Neuroscience, 3(9), Article 9. 10.1038/nrn920

Cohen, J. E., Holsen, L. M., Ironside, M., Moser, A. D., Duda, J. M., Null, K. E., Perlo, S., Richards, C. E., Nascimento, N. F., Du, F., Zuo, C., Misra, M., Pizzagalli, D. A., & Goldstein, J. M. (2023). Neural response to stress differs by sex in young adulthood. Psychiatry Research: Neuroimaging, 332, 111646. 10.1016/j.pscychresns.2023.111646

Cohen Kadosh, K., Krause, B., King, A. J., Near, J., & Cohen Kadosh, R. (2015). Linking GABA and glutamate levels to cognitive skill acquisition during development. Human Brain Mapping, 36(11), 4334–4345. 10.1002/hbm.22921

Delli Pizzi, S., Padulo, C., Brancucci, A., Bubbico, G., Edden, R. A., Ferretti, A., Franciotti, R., Manippa, V., Marzoli, D., Onofrj, M., Sepede, G., Tartaro, A., Tommasi, L., Puglisi-Allegra, S., & Bonanni, L. (2016). GABA content within the ventromedial prefrontal cortex is related to trait anxiety. Social Cognitive and Affective Neuroscience, 11(5), 758–766. 10.1093/scan/nsv155

Elliott, K. A. C. (1965). γ-aminobutyric acid and other inhibitory substances. British Medical Bulletin, 21(1), 70–75. 10.1093/oxfordjournals.bmb.a070360

Etkin, A., Büchel, C., & Gross, J. J. (2015). The neural bases of emotion regulation. Nature Reviews Neuroscience, 16(11), Article 11. 10.1038/nrn4044

Fair, D. A., Cohen, A. L., Dosenbach, N. U. F., Church, J. A., Miezin, F. M., Barch, D. M., Raichle, M. E., Petersen, S. E., & Schlaggar, B. L. (2008). The maturing architecture of the brain’s default network. Proceedings of the National Academy of Sciences, 105(10), 4028–4032. 10.1073/pnas.0800376105

Hasler, G., Buchmann, A., Haynes, M., Müller, S. T., Ghisleni, C., Brechbühl, S., & Tuura, R. (2019). Association between prefrontal glutamine levels and neuroticism determined using proton magnetic resonance spectroscopy. Translational Psychiatry, 9(1), Article 1. 10.1038/s41398-019-0500-z

Hasler, G., van der Veen, J. W., Geraci, M., Shen, J., Pine, D., & Drevets, W. C. (2009). Prefrontal Cortical Gamma-Aminobutyric Acid Levels in Panic Disorder Determined by Proton Magnetic Resonance Spectroscopy. Biological Psychiatry, 65(3), 273–275. 10.1016/j.biopsych.2008.06.023

Huang, Z. J., Di Cristo, G., & Ango, F. (2007). Development of GABA innervation in the cerebral and cerebellar cortices. Nature Reviews Neuroscience, 8(9), Article 9. 10.1038/nrn2188

Khona, M., & Fiete, I. R. (2022). Attractor and integrator networks in the brain. Nature Reviews Neuroscience, 23(12), Article 12. 10.1038/s41583-022-00642-0

Koh, W., Kwak, H., Cheong, E., & Lee, C. J. (2023). GABA tone regulation and its cognitive functions in the brain. Nature Reviews Neuroscience, 1–17. 10.1038/s41583-023-00724-7

Kovacs, M. (1992). Children’s Depression Inventory. Multi Health Systemes Inc.

La Greca, A. M. (1999). Manual for the Social Anxiety Scales for Children and Adolescents—Revised. University of Miami.

Larsen, B., Cui, Z., Adebimpe, A., Pines, A., Alexander-Bloch, A., Bertolero, M., Calkins, M. E., Gur, R. E., Gur, R. C., Mahadevan, A. S., Moore, T. M., Roalf, D. R., Seidlitz, J., Sydnor, V. J., Wolf, D. H., & Satterthwaite, T. D. (2022). A developmental reduction of the excitation:inhibition ratio in association cortex during adolescence. Science Advances, 8(5), eabj8750. 10.1126/sciadv.abj8750

LeDuke, D. O., Borio, M., Miranda, R., & Tye, K. M. (2023). Anxiety and depression: A top-down, bottom-up model of circuit function. Annals of the New York Academy of Sciences, 1525(1), 70–87. 10.1111/nyas.14997

Lee, M. D., & Wagenmakers, E.-J. (2014). Bayesian Cognitive Modeling: A Practical Course. Cambridge University Press.

Levelt, C. N., & Hübener, M. (2012). Critical-Period Plasticity in the Visual Cortex. Annual Review of Neuroscience, 35(1), 309–330. 10.1146/annurev-neuro-061010-113813

Li, H., Heise, K.-F., Chalavi, S., Puts, N. A. J., Edden, R. A. E., & Swinnen, S. P. (2022). The role of MRS-assessed GABA in human behavioral performance. Progress in Neurobiology, 212, 102247. 10.1016/j.pneurobio.2022.102247

Long, Z., Medlock, C., Dzemidzic, M., Shin, Y.-W., Goddard, A. W., & Dydak, U. (2013). Decreased GABA levels in anterior cingulate cortex/medial prefrontal cortex in panic disorder. Progress in Neuro-Psychopharmacology & Biological Psychiatry, 44, 131–135. 10.1016/j.pnpbp.2013.01.020

Lydiard, R. B. (2003). The role of GABA in anxiety disorders. Journal of Clinical Psychiatry, 64(SUPPL. 3), 21–27. Scopus.

Makowski, D., Ben-Shachar, M. S., Patil, I., & Lüdecke, D. (2020). Methods and Algorithms for Correlation Analysis in R. Journal of Open Source Software, 5(51), 2306. 10.21105/joss.02306

Makowski, D., Wiernik, B. M., Patil, I., Lüdecke, D., Ben-Shachar, M. S., White, M., & Rabe, M. M. (2023). correlation: Methods for Correlation Analysis (0.8.4) [Computer software]. https://cran.r-project.org/web/packages/correlation/index.html

Mauss, I. B., Bunge, S. A., & Gross, J. J. (2007). Automatic Emotion Regulation. Social and Personality Psychology Compass, 1(1), 146–167. 10.1111/j.1751-9004.2007.00005.x

Millan, M. J. (2003). The neurobiology and control of anxious states. Progress in Neurobiology, 70(2), 83–244. 10.1016/s0301-0082(03)00087-x

Mlynárik, V., Gambarota, G., Frenkel, H., & Gruetter, R. (2006). Localized short-echo-time proton MR spectroscopy with full signal-intensity acquisition. Magnetic Resonance in Medicine, 56(5), 965–970. 10.1002/mrm.21043

Owens, D. F., & Kriegstein, A. R. (2002). Is there more to GABA than synaptic inhibition? Nature Reviews Neuroscience, 3(9), 715–727. Scopus. 10.1038/nrn919

Page, C. E., & Coutellier, L. (2019). Prefrontal excitatory/inhibitory balance in stress and emotional disorders: Evidence for over-inhibition. Neuroscience & Biobehavioral Reviews, 105, 39–51. 10.1016/j.neubiorev.2019.07.024

Perica, M. I., Calabro, F. J., Larsen, B., Foran, W., Yushmanov, V. E., Hetherington, H., Tervo-Clemmens, B., Moon, C.-H., & Luna, B. (2022). Development of frontal GABA and glutamate supports excitation/inhibition balance from adolescence into adulthood. Progress in Neurobiology, 219, 102370. 10.1016/j.pneurobio.2022.102370

Porges, E. C., Jensen, G., Foster, B., Edden, R. A. E., & Puts, N. A. J. (2021). The trajectory of cortical gaba across the lifespan, an individual participant data meta-analysis of edited mrs studies. eLife, 10. Scopus. 10.7554/eLife.62575

Provencher, S. W. (1993). Estimation of metabolite concentrations from localizedin vivo proton NMR spectra. Magnetic Resonance in Medicine, 30(6), 672–679. 10.1002/mrm.1910300604

R Core Team. (2023). R: A Language and environment for statistical computing. (4.3.0) [Computer software]. R Foundation for Statistical Computing. https://www.R-project.org/

Simpson, R., Devenyi, G. A., Jezzard, P., Hennessy, T. J., & Near, J. (2017). Advanced processing and simulation of MRS data using the FID appliance (FID-A)—An open source, MATLAB-based toolkit. Magnetic Resonance in Medicine, 77(1), 23–33. 10.1002/mrm.26091

Spielberger, C. D. (1973). Manual for the state trait anxiety inventory for children. Consulting Psychologists Press. https://www.scirp.org/(S(i43dyn45teexjx455qlt3d2q))/reference/ReferencesPapers.aspx?ReferenceID=720680

Spielberger, C. D., Gorsuch, R. L., Lushene, R., Vagg, P. R., & Jacobs, G. A. (1983). State-Trait Anxiety Inventory for Adults. Mind Garden Inc.

Stanley, J. A., Burgess, A., Khatib, D., Ramaseshan, K., Arshad, M., Wu, H., & Diwadkar, V. A. (2017). Functional dynamics of hippocampal glutamate during associative learning assessed with in vivo 1H functional magnetic resonance spectroscopy. NeuroImage, 153, 189–197. 10.1016/j.neuroimage.2017.03.051

Strasser, A., Xin, L., Gruetter, R., & Sandi, C. (2019). Nucleus accumbens neurochemistry in human anxiety: A 7 T 1H-MRS study. European Neuropsychopharmacology, 29(3), 365–375. 10.1016/j.euroneuro.2018.12.015

Takado, Y., Takuwa, H., Sampei, K., Urushihata, T., Takahashi, M., Shimojo, M., Uchida, S., Nitta, N., Shibata, S., Nagashima, K., Ochi, Y., Ono, M., Maeda, J., Tomita, Y., Sahara, N., Near, J., Aoki, I., Shibata, K., & Higuchi, M. (2022). MRS-measured glutamate versus GABA reflects excitatory versus inhibitory neural activities in awake mice. Journal of Cerebral Blood Flow & Metabolism, 42(1), 197–212. 10.1177/0271678X211045449

Thompson, R. A., Lewis, M. D., & Calkins, S. D. (2008). Reassessing Emotion Regulation. Child Development Perspectives, 2(3), 124–131. 10.1111/j.1750-8606.2008.00054.x

Uhlhaas, P. J., Davey, C. G., Mehta, U. M., Shah, J., Torous, J., Allen, N. B., Avenevoli, S., Bella-Awusah, T., Chanen, A., Chen, E. Y. H., Correll, C. U., Do, K. Q., Fisher, H. L., Frangou, S., Hickie, I. B., Keshavan, M. S., Konrad, K., Lee, F. S., Liu, C. H., … Wood, S. J. (2023). Towards a youth mental health paradigm: A perspective and roadmap. Molecular Psychiatry, 1–11. 10.1038/s41380-023-02202-z

Werker, J. F., & Hensch, T. K. (2015). Critical Periods in Speech Perception: New Directions. Annual Review of Psychology, 66(1), 173–196. 10.1146/annurev-psych-010814-015104

Zacharopoulos, G., Sella, F., & Cohen Kadosh, R. (2021). The impact of a lack of mathematical education on brain development and future attainment. Proceedings of the National Academy of Sciences, 118(24), e2013155118. 10.1073/pnas.2013155118

